# Interventions to prevent neonatal healthcare-associated infections in low-resource settings: a systematic review

**DOI:** 10.1101/2020.12.17.20248047

**Authors:** Felicity C. Fitzgerald, Walter Zingg, Gwendoline Chimhini, Simbarashe Chimhuya, Stefanie Wittmann, Helen Brotherton, Ioana D. Olaru, Samuel R. Neal, Neal Russell, André Ricardo Araujo da Silva, Mike Sharland, Anna C Seale, Mark F Cotton, Susan Coffin, Angela Dramowski

## Abstract

**Background:** Clinically suspected and laboratory-confirmed bloodstream infections are frequent causes of morbidity and mortality during neonatal care. The most effective infection prevention and control (IPC) interventions for neonates in low-and-middle-income countries (LMIC) are unknown.

**Aim:** To identify effective interventions in the prevention of hospital-acquired bloodstream infections in LMIC neonatal units.

**Methods:** Medline, PUBMED, The Cochrane Database of Systematic Reviews, EMBASE, and PsychInfo (January 2003 – October 2020) were searched to identify studies reporting single or bundled interventions for prevention of bloodstream infections in LMIC neonatal units.

**Results:** Our initial search identified 5206 articles; following application of filters, 27 publications met the inclusion and ICROMS assessment criteria and were summarised in the final analysis. No studies were carried out in low-income countries, only one in sub-Saharan Africa and just two in multiple countries. Of the 18 single intervention studies, most targeted skin (n=4) and gastrointestinal mucosal integrity (n=5). Whereas emollient therapy and lactoferrin achieved significant reductions in proven neonatal infection, glutamine and mixed probiotics showed no benefit. Chlorhexidine gluconate for cord care and kangaroo mother care reduced infection in individual single-centre studies. Of the nine studies evaluating bundles, most focused on prevention of device-associated infections and achieved significant reductions in catheter- and ventilator-associated infections.

**Conclusion:** There is a limited evidence-base for the effectiveness of IPC interventions in LMIC neonatal units; bundled interventions targeting device-associated infections were most effective. More multi-site studies with robust study designs are needed to inform IPC intervention strategies in low-resource neonatal units.

**Sources of Support:** FF is supported by the Academy of Medical Sciences, the funders of the Starter Grant for Clinical Lecturers scheme and UCL Great Ormond Street NIHR Biomedical Research Centre. AD is supported by the Fogarty International Center of the National Institutes of Health, Emerging Global Leader Award Number K43-TW010682.

## Introduction

The World Health Organization (WHO) estimates that bacterial infections cause ∼25% of the 2.8 million annual neonatal deaths and long-term neurodevelopmental disabilities in survivors ^1^. Hospital-acquired infection (HAI) is a major cause of neonatal morbidity and mortality with prevalence ratios in low and middle-income countries (LMICs) 3–20 times higher than high-income countries ^2^. Traditional definitions, applied in high-income countries, use a 72-hour cut-off to differentiate early-from late-onset infection: the former associated with vertical transmission of pathogens such as Group B Streptococcus, the latter with horizontal transmission of hospital-acquired pathogens, often associated with prematurity and invasive procedures such as intravenous catheterisation. However, particularly in LMICs, there is recognition that facility-based delivery is itself a risk for HAIs, with pathogens such as *Klebsiella pneumoniae* (previously associated with late-onset infection) commonly isolated in the first 24 hours of life^2, 3^. This observation informs the Strengthening the Reporting of Observational Studies in Epidemiology for Newborn Infection (STROBE-NI) guidelines, which recommend recording the timing of symptom onset rather than the binary early/late-onset dicohotomy^1^. It also raises questions about fundamental differences in the mechanisms of neonatal infections in LMICs, as compared to high-income countries. The leading neonatal pathogens are increasingly resistant to first and second-line antimicrobials, with substantial resistance to commonly used agents including ampicillin (89% of *Escherichia coli)*, ceftriaxone (49% of *Klebsiella* spp. Isolates) and cloxacillin (40% of *Staphylococcus aureus*)^3^.

In this context, effective, feasible and affordable interventions to enhance infection prevention and control (IPC) in LMIC neonatal units are critical to prevent both neonatal mortality, and emerging antimicrobial resistance. However, even in high-income settings, implementing effective prevention measures are challenging, and a robust evidence-base on what tools to use is limited. Randomised controlled trials are considered the gold-standard for generating evidence in general. However, best practice procedures and quality improvement interventions must be contextual for maximum impact. As interventions are seldom identical across trial sites, patient-level randomisation is often not possible. Trials within hospitals (randomizing wards for example) are at risk of bias due to movement between wards of staff and patients. Furthermore, matching hospitals for randomisation can be complex ^4^.

To address these methodological challenges, new study designs, such as interrupted time series for cohorts and hospital-level stepped-wedge cluster-randomisation have been adopted. In addition, qualitative research aiming at understanding behaviour change is increasingly used to complement quantitative data ^4^. For neonates in LMICs, various HAI prevention strategies have been suggested but only studied in small and single centre studies. To date, the evidence-base in these settings has not yet been systematically assessed. We set out to review a broad range of potential interventions (both single and bundled), aiming to reduce healthcare-associated infections, with a focus on bloodstream infections (BSIs) in LMIC neonatal units.

## Methods

This systematic review was conducted in adherence with the Preferred Reporting Items for Systematic reviews and Meta-Analyses (PRISMA) statements of evaluations of healthcare interventions^5^. We registered the search strategy on the international prospective register of systematic reviews (CRD42018112346 on PROSPERO, see supplementary files).

### Search strategy

We searched Medline, The Cochrane Database of Systematic Reviews, EMBASE, and PsychInfo (1 January 2003 – 31 October 2020) to identify studies reporting on the effectiveness of interventions to prevent infections in LMIC neonatal wards and neonatal intensive care units. We selected the year 2003 to reflect the rapid evolution and spread of resistant bacteria causing HAIs in the last 17 years. IPC interventions were defined as any intervention aiming to prevent the development of a healthcare-associated bacterial or fungal infection such as BSI, meningitis, laboratory-confirmed urinary tract infection, or clinically suspected but culture-negative infections.

We limited results by age (neonates 0-27 days or 0-89 days if admitted on a neonatal ward or NICU), by location (LMIC as defined by the 2021 World Bank classification^6^), by language (articles written in English, German, French, Italian, Portuguese and Spanish were included), and by relevant filters as per exclusion criteria (For a full list of terms and filters see supplementary files). Our primary outcome was the effect of the interventions on (a) incidence of infection, or (b) attributable mortality, depending on study definitions. Fungal or bacterial hospital-acquired invasive infections in hospitalised neonates were the primary events for study. Secondary outcomes included impact on incidence of laboratory-confirmed urinary tract infection, thrombophlebitis, necrotizing enterocolitis (NEC), device-associated infections (clinically suspected or culture proven) and clinically suspected infection where laboratory cultures were negative or not available.

### Inclusion criteria

Studies were eligible for full-text review if conducted in hospitalised neonates, including neonatal ward and/or NICU settings, with a detailed description of the intervention. We included both single interventions (e.g. probiotics, kangaroo mother care (KMC), breastfeeding, fluconazole prophylaxis) and bundled interventions (e.g. vascular device care, hand hygiene, and healthcare worker education combined). Studies conducted in several countries including both high- and low- or middle-income countries (as per World Bank 2021 regions) could be included if possible to extract data from the LMIC settings. Study designs included randomised controlled trials, controlled and non-controlled before-after, controlled and non-controlled interrupted time series and cohort studies.

### Exclusion criteria

We excluded letters, opinion articles and reviews that did not report primary data. IPC interventions conducted during maternal care, in community-based settings and during outbreaks were excluded. We also excluded studies conducted exclusively in high-income countries as per World Bank 2021 regions^6^. Interventions targeting viral infections (including HIV), infants older than 3 months, or involving vaccination, diagnostic tools, infection prediction scores were excluded. We also excluded studies addressing IPC interventions on mixed neonatal/paediatric populations where extraction of neonatal data was not possible; and where only abstracts were available despite contacting the corresponding author. Finally, we excluded studies where bacterial colonisation as opposed to invasive infection was the outcome, if bloodstream infection was not also included.

### Study selection process

The initial eligibility assessment of titles and abstracts identified by our search was conducted independently by FF and AD using the pre-determined inclusion and exclusion criteria. Disagreements on eligibility were resolved by consensus, if needed by consulting a third party. The reference lists of all eligible publications were screened for cross-referencing. After finalising articles for full-text review, two authors evaluated the quality of each eligible publication using the Integrated quality Criteria for the Review Of Multiple Study designs (ICROMS) tool^7^, with disagreements resolved as explained above. The ICROMS tool was designed to allow the systematic integration and assessment of differing study types including both quantitative and qualitative designs for reviews of public health interventions such as those targeting IPC ^7^. The ICROMS tool provides a list of quality criteria with a set of requirements specific for the study design. Studies are evaluated by a ‘decision matrix’ where mandatory criteria must be met. The robustness of the study is measured by a score (See Supplementary Tables for criteria and scoring). To pass to the final analysis, studies must meet the minimum score and the mandatory ICROMS criteria, after duplicate review.

### Data abstraction

We extracted data using a standardised data-collection form already independently piloted by FF and AD on a representative sample of studies. Study details collected on the form included: author/s, year of publication, country or countries where the study was performed, study design, study timeframe, setting (neonatal ward, NICU or both), intervention type, intervention details and effect. We grouped studies by intervention type: IPC bundles; catheter care; skin integrity and bacterial colonisation (umbilical cord care, skin cleansing, emollients and/or massage); fluconazole prophylaxis; hand hygiene; KMC; rooming-in/parental involvement in neonatal care; and gastrointestinal integrity (probiotics and feeding practices). Data synthesis involved the collation and tabulation of results by intervention type, summarizing the key intervention/s and their effectiveness in IPC for hospitalised neonates (using either relative risk, odds ratios or hazard ratios as reported by each study). We did not undertake a meta-analysis due to diversity of study type, interventions and outcomes i.e. although all studies targeted reduction of neonatal infections, each study had different modes of action for the intervention and/or major differences in study design that precluded combining data.

## Results

We identified 5206 articles on initial searching, after removal of duplicates (Figure 1). Filter application (see appendix) reduced this to 1799 titles and abstracts then reviewed independently by two study authors (FF and AD) for relevance. Of these, 124 were selected for full text review in duplicate and ICROMS scoring, leading to another 97 exclusions and 27 selected for inclusion in the final review (Tables 1 and 2). Forty studies were excluded for either missing mandatory ICROMS criteria or ICROMS scores below the cut-off for the particular study design. Of the included studies, 8 were conducted in lower middle-income countries and 19 in upper middle-income countries (only two studies were multi-country). None were conducted in low-income countries. Including multi-site studies and using the 2021 World Bank regions, 14 study sites were in Latin America/Caribbean, fourteen in South-East Asia/Pacific, five in the Middle East/North Africa, three in Europe/Central Asia and one in sub-Saharan Africa^6^. Eighteen studies evaluated single interventions and nine evaluated bundled interventions (two of which were conducted in multiple countries).

**Table 1:** Studies reaching ICROMS Criteria for inclusion describing single interventions for the prevention of hospital-acquired neonatal bloodstream infections and clinically-suspected infection in low-resource settings (January 2003 – January 2020)

| Author, year | Study design* | Country | Population/ Setting | Sample Size | Intervention type | Intervention | Outcome | Key findings |
| --- | --- | --- | --- | --- | --- | --- | --- | --- |
| Akin, 2014<sup>8</sup> | RCT | Turkey | Preterm <32 weeks gestation or <1500g birthweight neonates admitted to one NNU | 50 | Probiotics /feeding | Oral lactoferrin 200 mg/day versus placebo | Episodes of culture-proven nosocomial infection and NEC | Reduction in infection in intervention versus control: 4.4 vs. 17.3/1,000 patient days, p = 0.007.<br>No episodes of NEC in either group |
| Kaur, 2015<sup>9</sup> | RCT | India | Neonates <2000g birthweight admitted to one NNU | 130 | Probiotics/ feeding | Oral bovine lactoferrin versus placebo (80-142 mg/kg/day) | Incidence of first episode of culture-proven LOS (bacterial or fungal), probable infection, any LOS, infection-attributable mortality | Reduction in LOS in intervention versus placebo: 2/63 (3.2%) vs. 9/67(13.4%); risk ratio, 0.211; 95% CI, 0.044-1.019; p = 0.036.<br>Reduction of infection-attributable mortality in intervention: 0/63 (0%) vs. 5/67 (7.5%); p = 0.027. |
| Li, 2007<sup>11</sup> | RCT | China | Neonates <37 weeks gestation & <2000g birthweight admitted to one NNU | 53 | Probiotics/feeding | Parenteral glutamine supplementation versus none | Growth and development, tolerance to oral feeding; nosocomial infection | Non-significant reduction in nosocomial infection in intervention versus control : 10% versus 16%, p=0.518 |
| Ochoa, 2015<sup>10</sup> | RCT | Peru | Neonates 500g-2500g in three NNUs | 190 | Probiotics/feeding | Oral bovine lactoferrin (200mg/kg) versus placebo | Incidence of first episode of LOS (culture-proven or clinical), frequency of culture-proven LOS, incidence of NEC, length of stay, overall mortality, infection-related mortality, other adverse events, treatment intolerance | Infection incidence: 12/95 (12.6%) versus 21/95 (22.1%) in interventions versus control, p=0.085.<br>Subsequent subgroup analysis: significant reduction in infection in <1500g |
| Wang, 2014<sup>12</sup> | RCT | China | Term neonates admitted to NNU | 100 | Probiotics/feeding | Administration of mixed probiotic (*L. casei*, *L. acidophilus*, *Bacillus subtilis*, *E. faecalis*) versus placebo | Nosocomial pneumonia, nosocomial infection (culture-proven), multiple organ dysfunction syndrome, NEC, diarrhoea | Non-significant reduction in infection in intervention versus control: 4% versus 2%, p=0.4<br>Similar in NEC: 4% versus 8%, p=0.47<br>Significant reduction in nosocomial pneumonia: 16% versus 36%, p=0.023 |
| Darmstadt, 2004<sup>13</sup> | RCT | Egypt | 1 NICU, neonates <34 weeks GA and <72h of life | 103 | Emollient therapy | Sunflower seed oil (n=51) versus usual care (minimal use of emollients, n=52) | Incidence of culture-proven infection, skin condition score, mortality from infection | Significant reduction in nosocomial infections with sunflower oil versus controls (adjusted IRR, 0.46; 95% CI, 0.26-0.81; p = 0.007). No difference in mortality due to infection (adjusted odds ratio 0.72, 95% CI 0.39-1.34. Significantly improved skin condition scores in intervention group. |
| Darmstadt, 2005<sup>14</sup> | RCT | Bangladesh | 1 NNU, neonates <33 weeks GA and <72 hours of life | 497 | Emollient therapy | sunflower seed oil (n=159), Aquaphor (n=157), usual care (n= 181) | Incidence of culture-proven nosocomial BSI | Significant decrease in nosocomial infections with sunflower oil versus controls (adjusted IRR 0.59, 95% CI 0.37–0.96, p=0.032). Aquaphor: non-significant decrease (adjusted IRR 0.60, 0.35–1.03, p=0.065). |
| Erdemir, 2015<sup>16</sup> | RCT | Turkey | 1 NICU, neonates <34 weeks GA and ≤24 hours of life | 197 | Emollient therapy | Aquaphor emollient versus routine care (none) | Incidence of neonatal infection, skin colonisation, incidence of UTI | No difference in incidence of infection as a one-off outcome, (41/100 versus 43/97 intervention versus controls, p= 0.63) or culture -proven infection (23/100 (intervention group) versus 19/97 (controls), p=0.42) |
| Salam, 2015<sup>15</sup> | RCT | Pakistan | 1 NICU, neonates 26-37 weeks gestation | 258 | Emollient therapy | Daily topical application of coconut oil versus no intervention | Incidence of HAI, weight gain, skin condition, mortality at 28 days of life | Significant reduction in culture-proven infection in intervention versus controls (9/128 versus 27/130), adjusted hazard of HAI 6.0 (95% CI 2.3-16) in controls; incidence of HAI 40 versus 219/1000 patient days in intervention group versus controls. Improved weight gain and skin condition in the intervention group, no impact on mortality or duration of admission. |
| Gathwala, 2013<sup>17</sup> | RCT | India | 1 NICU, neonates $\leq 32$ GA $\leq 1500$g | 140 | Chlorhexidine gluconate for cord care | Daily application of 2.5% CHG (n=70) to the umbilical cord vs. 'dry' cord care (n=70) | Time to cord separation (primary outcome)<br>Incidence of culture-proven neonatal infection, probable neonatal infection, meningitis, umbilical infection (secondary outcomes) | Significantly fewer episodes of culture proven infection (2 versus 15, p= 0.02, absolute risk 21%, versus 3%, absolute risk reduction 19%, confidence intervals not shown), in interventions versus control; borderline significantly greater episodes of probable infection (10 versus 3, p=0.052) interventions versus controls. Significant reduction in time to cord separation in intervention group (mean 9 days versus 10 days, p=0.02). |
| Gupta, 2016<sup>18</sup> | RCT | India | 1 paediatric ward, neonates <24 hours of life | 140 | Chlorhexidine gluconate for skin cleansing | Daily application of 0.25% CHG to the whole body (n=70) versus tepid water (n=70) | Incidence of culture-proven HA-neonatal infection (cultures taken on days 1, 3 and 7 of life) | 6/168 (3.6%) blood culture samples positive in intervention group versus 12/175 (6.9%) in the controls, (p=0.195). |
| Boo, 2007<sup>19</sup> | RCT | Malaysia | Stable neonates <1500g birthweight admitted to one NNU | 126 | KMC | Intermittent skin to skin contact for minimum 1 hour/day (n=62) versus standard care (n=64) | Weight gain, occipitofrontal circumference, breast feeding. Infection and NEC as secondary outcomes | No significant difference in culture-proven infection: 2/64 neonates (intervention group) versus 1/64 (controls, p=1.0)<br>No episodes of NEC in either group. |
| Charpak, 1997<sup>20</sup> | RCT | Colombia | Stable, neonates, birthweight <2000g admitted to one NNU | 746 | KMC | Continuous KMC (n= 382) versus traditional management (n=364) | Growth and mortality to 40/41 weeks corrected gestational age. Severe infection requiring systemic antibiotics and nosocomial infections secondary outcomes | Similar numbers of infectious episodes 49/382 (intervention) versus 44/364 (controls), but more mild/moderate infectious episodes (7% interventions, versus 3%, controls), absolute figures not given. Reduction in nosocomial infections: 8% versus 4% in interventions/controls, p=0.026) absolute figures not given |
| Li, 2017<sup>23</sup> | NCBA | China | Stable Neonates >1500g in one NNU | 1446 | Rooming-in | Neonates moved to Room in from NICU (n=1018) versus those eligible to move but staying in NICU (n=428). 629 admitted directly to Room-in | Mortality, growth, duration of admission. Secondary outcomes: nosocomial infection and NEC (unclear how defined) | No difference in nosocomial infection: 100/1081 versus 48/428 in interventions versus controls, p=0.41; Fewer neonates in intervention group with NEC: 7/1081, versus 8/428, (p=0.04).<br>Reduction in mortality: 2% versus 0%, p<0.001 |
| Parikh, 2007<sup>25</sup> | RCT | India | Preterm neonates <1500g admitted to one tertiary NNU | 120 | Fluconazole prophylaxis | Fluconazole prophylaxis within first 3 days to day 28 or discharge/death if sooner (n=60) versus placebo (n=60) | Fungal colonisation and invasive fungal infection- cultures taken on days 1-3, 7, 14, 21 and 28 | No reduction in invasive candida infection detected by blood cultures: 16/60 episodes versus 15/60 in intervention versus control, p=0.835; Of note, 30/31 of invasive species were non-albicans species. |
| Barrera, 2010<sup>21</sup> | Cohort | Colombia | All neonates admitted to one NNU | 6655 | Hand hygiene | Introduction of alcohol based hand rub (ABHR) dispensers; initial education; daily surveillance, quarterly feedback | HAI, CLABSI, VAP and UTI as per CDC definitions | 1260 patients with HAI, 724/1,848 episodes confirmed by culture. Trend in reduction of Methicillin resistant *Staphylococcus aureus*, 2.2 to 0.6 infections/1000 patient days in from 2001-2005, -30%, p=0.001<br>No trend in reduction of *Acinetobacter baumannii* (0.6-0.2/1000 patient days, p value not given)<br>Significant increase in use of alcohol based hand rub |
| Mendes, 2008<sup>22</sup> | RCT | Brazil | 1 NICU, all neonates ≤32 weeks GA and 750-1500g | 104 | Massage therapy | Massage therapy (tactile-kinesthetic stimulation, n=52) versus no intervention (n=52) | Primary outcome: Length of NNU stay; secondary outcomes: weight gain, time to enteral feeds, time to oral feeds, incidence of LOS (clinically and blood culture confirmed), incidence of NEC (clinical and radiological confirmation) | Lower incidence LOS in intervention versus controls (5/46 versus 18/47, p=0.005); 8 versus 22 pathogens identified in cultures (unclear how many cultures had multiple pathogens). |
| Barria, 2007<sup>24</sup> | RCT | Chile | “high-risk” neonates admitted to one NNU | 74 | Intravenous catheterisation | Peripherally inserted central catheters (n=37) versus standard peripheral intravenous catheters (n=37) | Length of neonatal intensive care unit stay and incidence of infection and phlebitis. | No difference in incidence of suspected infection between groups: 14/37 versus 8/37, p=0.127<br>Or culture-proven infection: 1/37 versus 2/37, p=0.53<br>Reduction in phlebitis: 4/37 versus 15/37, p= 0.007<br>No difference in length of stay: median 20 versus 17 days in intervention/control groups, p=0.158 |

Abbreviations: ABHR: Alcohol based hand rub; BSI: blood stream infections; CA-BSI: catheter-associated blood stream infections; CBA: controlled before and after; CI: confidence intervals; CLABSI: Central line associated bloodstream infection; CVC: central venous catheters; HAI: hospital acquired infection; ITS: interrupted time series; IRR: incident rate ratio; KMC: Kangaroo Mother Care; LOS: late onset infection; NCBA: non-controlled before and after; NEC: necrotizing enterocolitis; NNU: Neonatal unit; RCT: Randomised controlled trial; UTI: urinary tract infection; VAP: ventilator-associated pneumonia.

**Table 2:** Bundled interventions for the prevention of hospital-acquired neonatal bloodstream infections and clinically-suspected infection in low-resource settings (January 2003 – September 2018)

| Author, year | Study design | Country | Population/ Setting | Sample Size | Bundle elements | Outcome/s | Key findings |
| --- | --- | --- | --- | --- | --- | --- | --- |
| Azab, 2015<sup>26</sup> | NCBA | Egypt | 1 NICU, all NICU admissions with duration of invasive ventilation > 48 hours | 62 vs 81 | VAP prevention bundle + routine IPC measures: head-of-bed elevation, hand hygiene, sterile suctioning, strict indications for Intubation, re-intubation and suctioning, ventilator circuit change if visibly soiled or malfunctioning, mouth care, daily evaluation for readiness for extubation, sedation vacations. | VAP episodes per 1000 mechanical ventilator days<br>Length of stay in NICU<br>Overall mortality | VAP rate reduced from 36.4 to 23 episodes/1000 mechanical ventilation (MV) days (RR 0.565, 95%CI 0.408-0.782, p=0.0006) and reduced MV days/case in the post-intervention period (21.50±7.6 days to 10.36 ±5.2 days; p=0.0001). Trend towards reduction in NICU length of stay (LOS) (23.9 ± 10.3 to 22.8 ± 9.6 days, p=0.56) and overall mortality (25 % to 17.3 %, p=0.215). |
| Gilbert, 2014<sup>32</sup> | NC ITS | Brazil | 5 NNUs, all admissions <1500g | 679 vs 563 | Nurse training package, including IPC measures | Mortality in VLBW neonates (primary outcome)<br>Incidence of late onset infection, NEC, and other secondary outcomes | Despite improvement in nurses' knowledge and practices, there was no change in survival (pre-training 80%, post 78.2%), severe ROP (1.6 vs. 2.8%), late onset infection (11.3 vs. 12.3 cases/1,000 infant days) or other outcomes. |
| Gill, 2009<sup>31</sup> | NCBA | Philippines | 2 NICUs, all admissions between 2003 and 2004. | phase 1 =925; phase 2 = 902 | Bundle with blood culture quality improvement, provision of alcohol handrub, infection and HH surveillance, education, | Proportion of neonates newly colonised with resistant pathogens.<br>Secondary outcomes | Rates of colonisation with drug-resistant pathogens and rates of infection did not change significantly. Staff hand hygiene compliance improved compared to the |
|  |  |  |  |  | case discussions, infection control checklists | included bacteremia rates, cumulative mortality in NICU, and hand-hygiene compliance rates | control period (NICU1: relative risk (RR), 1.3; 95%CI 1.1–1.5; NICU2: RR 1.6; 95%CI 1.4–2.0). Overall mortality decreased (NICU1: RR 0.5; 95%CI 0.4–0.6; NICU2: RR, 0.8; 95%CI 0.7–0.9). |
| Leng, 2016<sup>33</sup> | Cohort | China | 1 NNU, consecutive outborn neonates <1500g | 86 vs 86 | Hypothermia prevention bundle including standardised transport procedures, skilled transfer teams, process reviews with feedback | Axillary temperature on arrival (primary outcome). Description of rates of NEC, early and late onset neonatal infection | Mean delivery room and NICU admission temperatures rose from 35.5 to 36.1° C and from 34.6 to 36.2° C ($p < 0.01$), with significantly decreased mortality ($p < 0.02$). There was no difference in the incidence of NEC and infection following implementation of the intervention. |
| Mwananyanda, 2019<sup>34</sup> | Cohort | Zambia | All admissions to one NNU | 2669: 852 baseline, 268 implementation, 1549 intervention evaluation | IPC training, text message reminders, alcohol hand rub, enhanced environmental cleaning, and weekly bathing of neonates $\geq 1.5$ kg with 2% chlorhexidine gluconate | Mortality primary outcome, HAI, BSI secondary outcomes | Absolute mean monthly mortality reduction –9% (95% CI, –11 to –7); overall relative mortality risk reduction 21% (RR 0.79 [95% CI, 0.76–0.83])<br>Incidence rate ratio of suspected infection (0.48-0.65) & pathogen-identified (0.28-0.62) decreased for all weight groups except <1kg suspected infection (1.38, $p=0.53$. P values for others all $< 0.001$) |
| Resende, 2011<sup>30</sup> | NCBA | Brazil | All admissions to one NNU | 251 | Catheter bundle: surveillance, feedback of CA-BSI; education, training, posters, hand hygiene; full-barrier precautions during CVC insertion; chlorhexidine skin cleaning; avoiding femoral site; removing unnecessary catheters | BSI rates pre/post intervention | Reduction in culture-proven CA-BSI incidence pre/post intervention (32% versus 20%, 24 versus 15 per 1000 catheter days; $p=0.04$) |
| Rosenthal, 2012<sup>27</sup> | NCBA | Argentina, Colombia, El Salvador, India, Mexico, Morocco, Peru, Turkey Philippines, Tunisia | 10 NICUs, all admissions to NICU | 1237 vs 5592 | VAP bundle with active surveillance, HH, readiness to wean assessment, oral antiseptics, non-invasive ventilation, orotracheal intubation, management of ventilation circuits. | VAP rates | The VAP rate declined from 17.8/1,000 MV-days to 12.0/1,000 MV-days; RR 0.67, 95%CI 0.50-0.91; a 33% reduction in VAP rate. |
| Rosenthal, 2013<sup>29</sup> | NCBA | El Salvador, Mexico, Philippines, Tunisia | 4 NICUs, all admissions to NICU | 374 vs 1867 | CLABSI prevention bundle: IPC interventions; education; outcome + process surveillance, feedback of CLABSI rates, performance feedback of IPC practices. | CLABSI rates | CLABSI rate decreased by 55%, from 21.4/1,000 central line (CL)-days in phase 1 to 9.7/1,000 CL-days in phase 2 (rate ratio 0.45; 95%CI 0.33–0.63). |
| Zhou 2013<sup>28</sup> | NCBA | China | 1 NICU, all admissions with duration of invasive ventilation >48 hours and at least 5 days NICU stay | 106 vs 169 vs 216 | Bundle: HH, waste disposal, patient isolation, ventilator disinfection, education, rational antibiotic use | VAP rates<br>Overall mortality | VAP rate decreased from 48.84/1,000 MV-days to 25.73/1,000 MV-days in phase 2 and 18.50/1,000 MV-days in phase 3 (P< .001). Overall mortality rate decreased from 14.0% in phase 1 to 2.9% in phase 2 and 2.7% in phase 3 (P < .000). |

Study design abbreviations: RCT: Randomised controlled trial; NCBA: non-controlled before and after; CBA: controlled before and after; ITS: interrupted time series. NNU: Neonatal unit; NICU: Neonatal intensive care unit; VAP = ventilator-associated pneumonia; MV= mechanical ventilation; RR = relative risk; NEC = necrotizing enterocolitis; HH = hand hygiene; IPC = infection prevention and control; CA-BSI: catheter associated blood stream infections; CVC: central venous catheters; BSI: blood stream infections; ABHR: Alcohol based hand rub; KMC: Kangaroo Mother Care; NEC: necrotizing enterocolitis; LOS: late onset infection.

**Figure 1.**
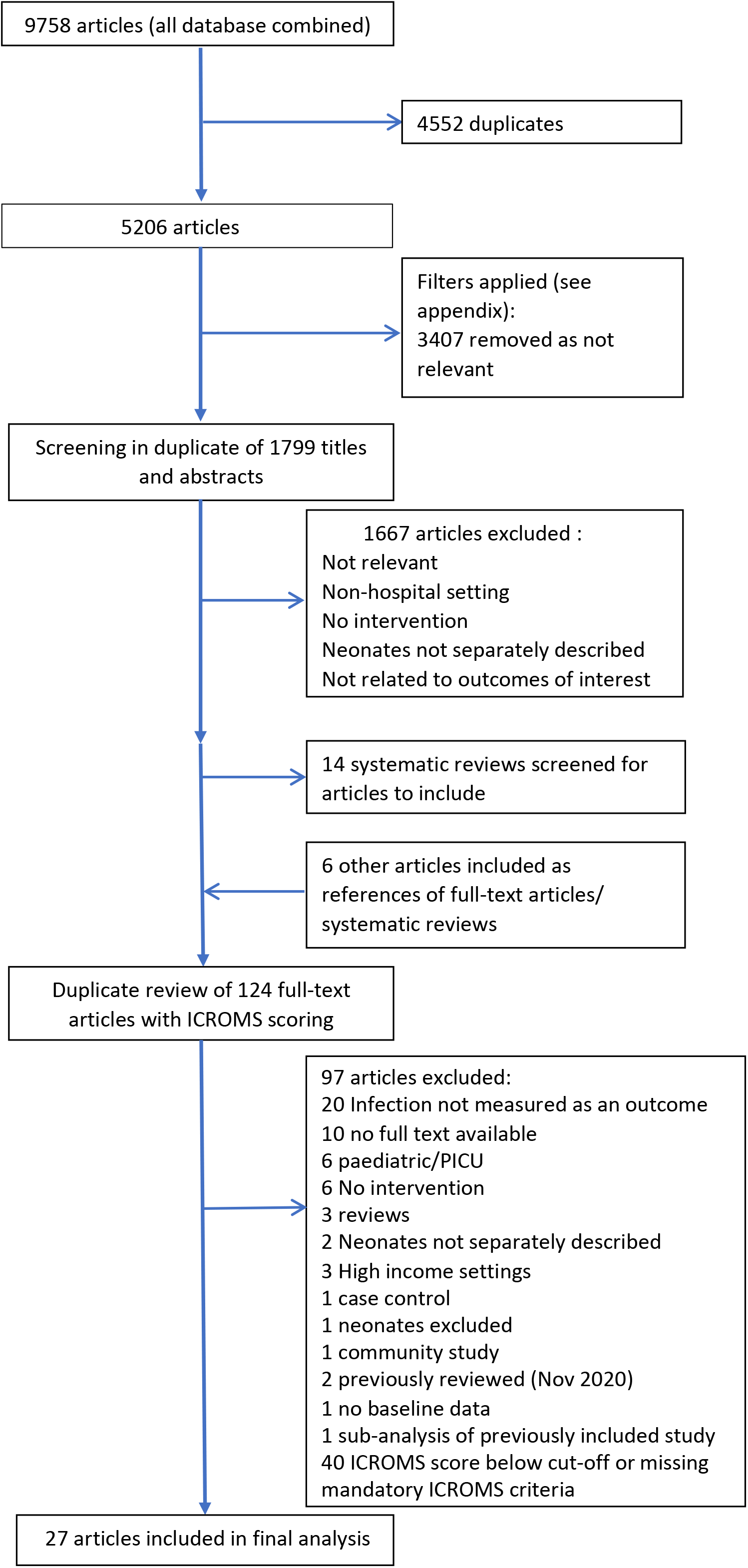
Search strategy for the identification and selection of publications reporting the effectiveness of interventions to prevent infections in neonatal wards and intensive care (January 2003-October 2020)

### Single Intervention studies

Of the single interventions (Table 1), probiotics/feeding interventions were the most commonly evaluated (five), followed by emollients (four), chlorhexidine cord cleansing (two) and KMC (two).

Three of the 5 probiotic/feeding interventions evaluated oral bovine lactoferrin versus placebo in a total of 370 neonates with birthweights <2500g ^8-10^. Varying bovine lactoferrin dosage (from 80-200mg/kg/day) and weight/gestational age thresholds made data incomparable and meta-analysis inappropriate. Two studies showed reduction in HAI in the intervention groups, one documenting 4.4 infections per 1000 patient days in the intervention arm versus 17.3 (p=0.007), the other finding a risk ratio of 0.211; (95% confidence intervals [CI] 0.044-1.019; p = 0.036) in those receiving the intervention versus placebo^8,9^. Two studies evaluated enteral supplements but neither reduced infection incidence (parenteral glutamine supplementation[p=0.518]^11^ nor mixed probiotic administration[p=0.4] ^12^).

For emollients, one group conducted two studies using sunflower seed oil in 103 Egyptian and 497 Bangladeshi neonates <72 hours of age, born at <34 or <33 weeks’ gestational age respectively^13, 14^. Both studies found that sunflower seed oil massage was associated with a significant decrease in the adjusted incidence rate ratio (aIRR, adjusted for weight on admission, gestational age and sex) of culture proven BSI than control (aIRR 0.46; 95% CI, 0.26–0.81; and 0.59, 95% CI 0.37-0.96). Notably, the Bangladeshi study showed no difference in the rate of clinically suspected infection triggering taking of blood cultures or antibiotic treatment rates between groups, although culture-proven BSI decreased in the intervention arm. Topical coconut oil was used in a Pakistani study in 270 neonates (26-34 weeks gestational age), first in the neonatal unit (NNU) and then at home^15^. Neonates randomised to the control arm had an increased risk of hospital-acquired BSI (adjusted hazard ratio 6.0, 95% CI 2.3-16). A Turkish study of 197 preterm neonates (<34 weeks’ gestation and <24 hours old) found no difference in mortality, incidence of culture-proven or clinically-suspected infection in patients randomised to receive Aquaphor emollient versus standard skin care^16^.

Two studies from India examined the impact of topical application of chlorhexidine gluconate; one in 140 neonates ≥32 weeks’ gestational age and ≥1500g using chlorhexidine 2.5% to clean the umbilical stump; the other in 140 neonates comparing whole body cleansing with chlorhexidine 0.25% versus tepid water^17, 18^. The first demonstrated a significant decrease in culture-proven BSI with chlorhexidine cord care (2 versus 15, p= 0.02, absolute risk 21%, versus 3%, absolute risk reduction 19%, confidence intervals not shown), although clinically-suspected infections increased in intervention versus control subjects (Table 1)^17^. The second study found a non-significant decrease in blood culture positivity with whole body cleansing (6/168 blood cultures positive in intervention group versus 12/175, p=0.195), possibly owing to a small sample size and that blood cultures were taken at set intervals regardless of clinical indication^18^.

Studies on KMC were carried out in Colombia and Malaysia, in 746 neonates <2000g and 126 neonates <1500g respectively^19, 20^. These studies evaluated substantially different KMC interventions (∼24 hours per day of KMC versus ≥1 hour per day of KMC (Table 1). The Colombian study found similar numbers of infectious episodes 49/382 (intervention) versus 44/364 (controls), although they describe a milder phenotype in the intervention arm, and a reduction in nosocomial infections (8% versus 4% in interventions/controls, p=0.026, absolute figures not given), without a clear distinction of the definition of ‘nosocomial’ versus other infections. In the Malaysian study, there were 2/64 infections in the intervention group versus 1/64 (controls, p=1.0).

A large cohort study in Colombia (6655 neonates) evaluating a hand hygiene intervention (alcohol-based handrub dispensers, daily surveillance and quarterly feedback), found a decreased incidence density of neonatal methicillin-resistant *Staphylococcus aureus* bloodstream infections (from 2.2 to 0.6 per 1000 patient days, p=0.01), although no decrease in *Acinetobacter baumannii*^21^ (0.6-0.2 per 1000 patient days, p not given).

A small Brazilian study of massage therapy versus no intervention (n=104) reported lower incidence of late onset infections in the intervention versus control groups^22^.

No study evaluating ‘rooming-in’ (defined as continuous presence of parent caregivers in the neonatal unit^23^), peripherally inserted central catheters versus standard intravenous catheters^24^ and fluconazole prophylaxis^25^ found differences in infection rates between the study arms (Table 1).

### Bundled interventions

Five of the 9 studies reporting the impact of IPC bundles (Table 2) focused on preventing device-associated infection^26-30^. One small, single-centre study in an Egyptian NICU, achieved significant reduction in ventilator-associated pneumonia (VAP) rates and mechanical ventilation days, with a trend towards reduction in NICU length of stay and overall mortality^26^. A multi-country study in 10 NICUs demonstrated significant reduction in VAP rates (RR 0.67, 95%CI 0.50-0.91), after implementation of a multi-modal strategy including hand hygiene, oral antiseptics, ventilator circuit management and enhanced infection surveillance^27^. A tertiary hospital, 50-bed NICU in China significantly reduced VAP rates, as well as overall mortality following implementation of a bundle including hand hygiene, ventilator disinfection, education and rational antibiotic use^28^.

Two studies targeted prevention of central line-associated bloodstream infection (CLABSI). A multi-country study in 4 NICUs demonstrated significant reduction in CLABSI rates following a multi-modal intervention strategy including education, enhanced process and outcome surveillance and staff feedback (rate ratio 0.45; 95%CI 0.33–0.63)^29^. A single-center Brazilian NICU significantly reduced CLABSI rates (24 versus 15 per 1000 catheter days; p=0.04) following implementation of a bundle including education, hand hygiene, CHG skin preparation and removal of unnecessary catheters^30^.

The first of two studies utilizing education/training interventions was a non-controlled ‘before after’ study conducted in 2 NICUs in the Philippines. The bundle focused on quality improvement in blood culture collection, hand hygiene compliance, use of infection control checklists and staff education. Although there was no change in the primary outcome (proportion of neonates newly colonised with resistant pathogens) or in the secondary outcome of bacteraemia, the study achieved improved hand hygiene compliance rates and reduction in overall mortality^31^. A Brazilian study in 5 neonatal units conducted an interrupted time-series analysis following introduction of a nurse training package including IPC measures. Despite improvement in nurses’ knowledge and practices, there was no change in mortality or rates of hospital-acquired BSI (11.3 vs. 12.3 cases/1,000 infant days)^32^.

A single-centre cohort study at a large, academic centre NICU in China enrolled outborn neonates <1500g to assess the impact of a hypothermia prevention bundle on admission temperature, rates of NEC and neonatal infection. Mean axillary temperature on arrival increased and overall mortality rates decreased significantly, however there was no difference in either NEC or infection incidence following the intervention^33^.

A recent, large cohort study in a Zambian neonatal unit evaluated the impact of IPC training, text message reminders for staff, hand hygiene promotion with alcohol-based handrub, enhanced environmental cleaning and weekly whole-body bathing of neonates ≥1.5 kg with 2% chlorhexidine gluconate. The bundle achieved significant reduction in overall mortality, clinically-suspected infection and culture-proven BSI for all birth weight groups except those <1kg.^34^ In a subsequent sub-analysis of the intervention group data, CHG bathing reduced the hazard rate of bloodstream infection among inborn babies ≥1.5 kg by a factor of 0.58 (p = 0.10, 95% CI: 0.31, 1.11)^35^.

## Discussion

Although infection is the most frequent complication of hospitalisation in LMIC neonates, the most effective IPC interventions remain unknown. We therefore conducted a systematic review of published studies describing the impact of various IPC interventions on healthcare-associated infection rates in LMIC NNUs. We identified 27 eligible publications that assessed single (n=18) and bundled IPC interventions (n=9). None were carried out in low-income countries, only one in sub-Saharan Africa and just two had sites in multiple countries. We found considerable heterogeneity of study design, analysis and outcomes selected, as well as diversity in the modes of infection prevention targeted (skin and gastrointestinal mucosal integrity, promotion of normal flora acquisition, reduction of bacterial pathogen colonisation). The evidence-base we have identified for the effectiveness of IPC interventions in LMIC neonatal units is limited, but appears most promising for bundled interventions targeting device-associated infections.

Limitations of this review include the paucity of published research on neonatal IPC from LMIC, the lack of multi-centre studies or large sample sizes and the failure to use optimal study interventional study designs. Although we endeavoured to be as inclusive as possible in our search terms, we only searched four databases and in six languages, so it is possible that we missed some relevant studies. It was not appropriate to do meta-analyses due to heterogeneity of both interventions and outcomes. Most studies were carried out in tertiary or academic neonatal units, which further limits the generalisability of the findings. Of note, although our initial search captured a large number of potentially eligible studies, full text review led to 40/120 (33%) papers being excluded due to not including mandatory criteria required by ICROMS, or having a low score for study design/analysis quality. Thus some geographical areas were not well represented-in particular sub-Saharan Africa with only one study included^34^. This highlights the challenges for clinicians in LMIC settings to identify and implement contextually appropriate evidence-based guidelines. It also demonstrates the difficulties of designing and analysing high quality IPC studies where facility, laboratory and statistical support may be lacking.

IPC studies are notoriously complex to design and implement, with issues of contamination between arms, the need for large scale randomisation (e.g. cluster randomisation of hospitals) and use of study designs unfamiliar to many academic clinicians e.g. interrupted time series analysis. IPC interventions also frequently involve behaviour change, which does not lend itself to RCT evaluation. In recognition of the importance of evaluating effective behaviour change in interventions in fields such as IPC, the UK Medical Research Council has developed guidance on how these studies should be designed and implemented^36^. Similarly the ICROMS score was developed to allow the inclusion of studies such as controlled before -after, non-controlled before-after studies and qualitative studies in assessing evidence, the exclusion of which from standard systematic reviews undermines their potential contribution to the evidence base^7^.

A major challenge in selecting the primary endpoint for neonatal IPC studies is the very low yield of blood cultures (the current gold standard for confirmation of BSI) in both high- and low-income settings. This necessitates recruitment of large numbers of neonates to conclusively demonstrate an intervention’s impact, which is often particularly challenging in LMIC owing to budgetary and logistic constraints. Sensitive and specific neonatal infection diagnostic tools that are accessible and affordable in LMIC settings are needed. In addition, standardised and validated definitions for clinically-suspected, culture-negative neonatal infections are required, to allow for comparison of findings across study sites. Use of multiple study outcomes (proven infection, clinically-suspected infection and mortality) may complicate interpretation of findings, particularly where the results are discrepant^14^. Until there is consensus on definitions of clinically-suspected neonatal infection, particularly in settings where cultures have limited availability, the issue of quantifying reduction in infection rates will persist.

Despite these inherent limitations in the available data, endpoint definitions and study methodologies used, we have conducted the first systematic review of IPC interventions for LMIC NNUs. We used a robust search strategy, long inclusion timeframe and ICROMS quality assessment to ensure we have identified all relevant and rigorously conducted research on this topic.

Among the single intervention studies, emollient therapy (sunflower oil) in low birthweight babies had the strongest evidence supporting its use, demonstrating reduced healthcare-associated infection rates in both studies^13, 14^. There was also evidence to support the use of oral bovine lactoferrin, although the studies were small and there was inconsistency in dosage used. This finding is echoed in a recent Cochrane review of studies in high and low-resource settings which concluded there was low-certainty evidence that lactoferrin supplementation could reduce late-onset sepsis, though not necrotising enterocolitis or all cause mortality^37^. Contrary to another previous Cochrane review, we did not find strong evidence for KMC as an intervention to reduce BSI in LMICs- only two studies fulfilled ICROMS criteria and only one had some evidence of impact on BSI^20, 38^. For studies that analyzed the impact of bundled interventions, the strongest evidence was generated from studies aiming to prevent device-associated infection. Bundles incorporating other interventions (education, infection surveillance with feedback, hand hygiene promotion and chlorhexidine gluconate bathing) were also effective, but the evidence was generated from single-centre or small studies.

Particular areas that appear promising for future research on neonatal IPC in LMIC are the use of chlorhexidine gluconate body washing and/or emollient therapy. Bundles that target neonatal BSI (the most common neonatal HAI) should be developed, utilising lessons learned from the success of bundles targeting device-associated infections. The ideal bundled intervention should target all portals of entry for pathogenic bacteria causing neonatal BSI. It could include avoidance of hospitalisation and/or invasive procedures, promotion of mucosal integrity (gut and skin), promotion of colonisation with normal flora and reduced colonisation with pathogenic bacteria.

Future studies in LMIC should utilise multi-national collaborations, standardise definitions (or at least clearly elucidate what criteria have been used) and use robust study designs e.g. individual randomised or cluster-randomised controlled trials and interrupted time-series analysis to generate evidence for IPC interventions that can be adopted in neonatal practice. Wherever possible guidelines such as Strengthening the Reporting of Observational Studies in Epidemiology for Newborn Infection (STROBE-NI) should be followed to allow for future comparisons between studies^1^.

## Conclusion

There is a limited evidence-base for IPC interventions in LMIC neonatal units. Overall, bundled interventions targeting prevention of device-associated infection are supported by the strongest evidence to date. More multi-site studies using standardised neonatal infection definitions and robust study designs are needed to inform IPC interventions for use in low-resource neonatal units.

## Data Availability

Systematic Review of Previously published data

Supplementary Appendix

Filters applied to initial search results with number of records identified

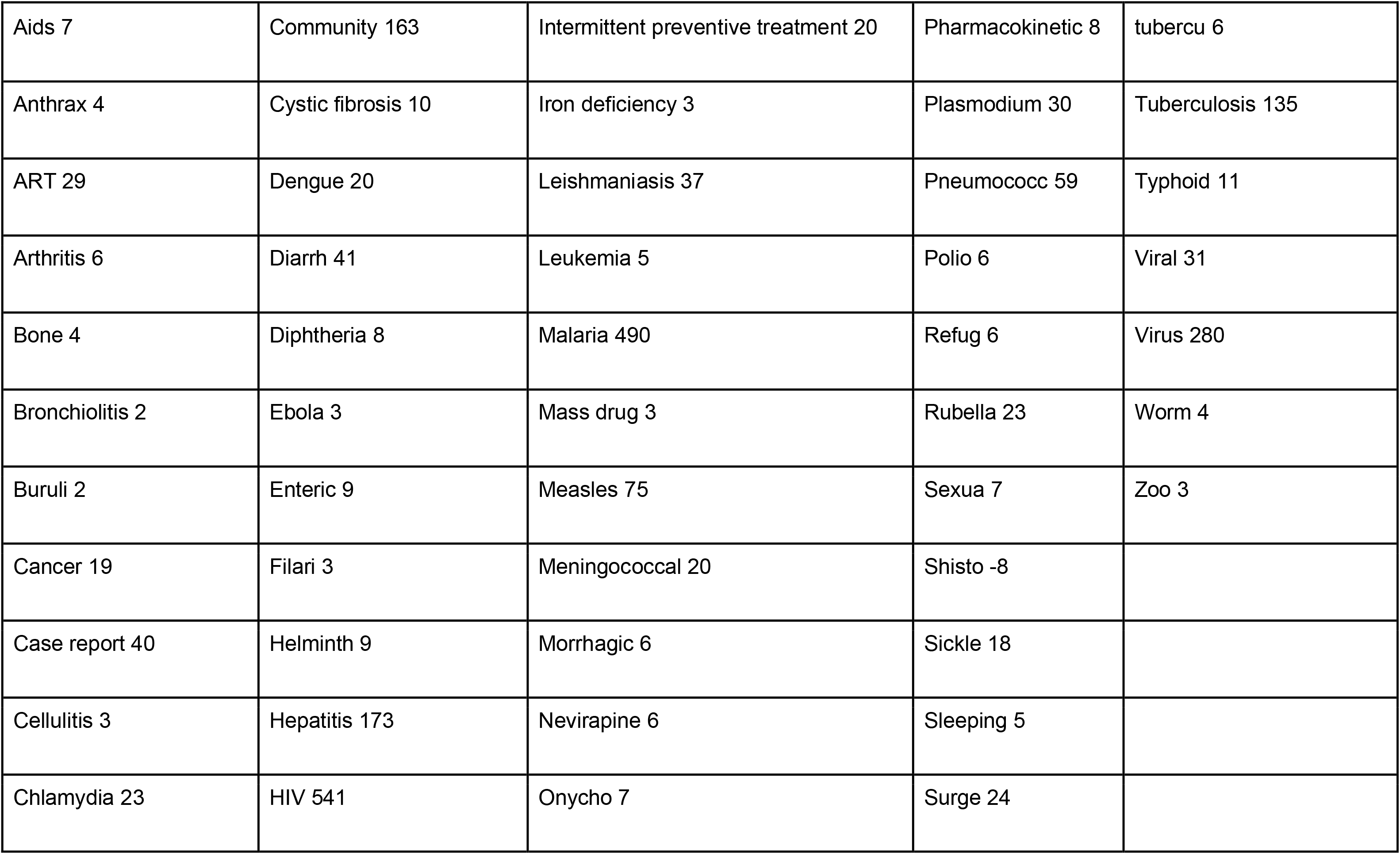
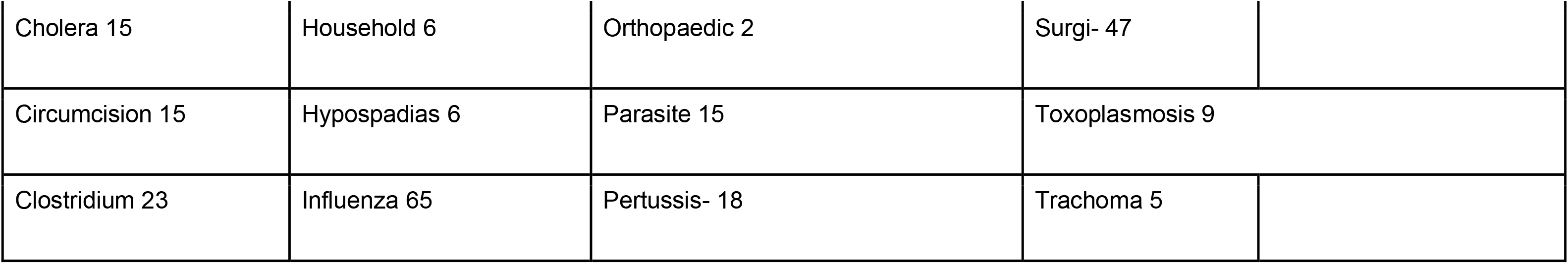

|  |  |  |  |  |
| --- | --- | --- | --- | --- |
| Aids 7 | Community 163 | Intermittent preventive treatment 20 | Pharmacokinetic 8 | tubercu 6 |
| Anthrax 4 | Cystic fibrosis 10 | Iron deficiency 3 | Plasmodium 30 | Tuberculosis 135 |
| ART 29 | Dengue 20 | Leishmaniasis 37 | Pneumococc 59 | Typhoid 11 |
| Arthritis 6 | Diarrh 41 | Leukemia 5 | Polio 6 | Viral 31 |
| Bone 4 | Diphtheria 8 | Malaria 490 | Refug 6 | Virus 280 |
| Bronchiolitis 2 | Ebola 3 | Mass drug 3 | Rubella 23 | Worm 4 |
| Buruli 2 | Enteric 9 | Measles 75 | Sexua 7 | Zoo 3 |
| Cancer 19 | Filari 3 | Meningococcal 20 | Shisto -8 |  |
| Case report 40 | Helminth 9 | Morrhagic 6 | Sickle 18 |  |
| Cellulitis 3 | Hepatitis 173 | Nevirapine 6 | Sleeping 5 |  |
| Chlamydia 23 | HIV 541 | Onycho 7 | Surge 24 |  |

|  |  |  |  |
| --- | --- | --- | --- |
| Cholera 15 | Household 6 | Orthopaedic 2 | Surgi- 47 |
| Circumcision 15 | Hypospadias 6 | Parasite 15 | Toxoplasmosis 9 |
| Clostridium 23 | Influenza 65 | Pertussis- 18 | Trachoma 5 |

---

**Supplementary Table 1:** ICROMS Quality Criteria applied for each study, by study design (from Zingg *et al*., 2016)^1^

| Quality Criteria |  | Study design |  |  |  |  |  |  |
| --- | --- | --- | --- | --- | --- | --- | --- | --- |
| Dimension | Specific criteria | RCT | CBA | CITS | NCITS | NCBA | CS | QUAL |
| 1. Clear aims and justification | a. Clear statement of aims | ++ | ++ | ++ | ++ | ++ | ++ | ++ |
|  | b. Rationale for number of pre- and post-intervention points or adequate baseline measurement |  |  | + | ++ | ++ |  |  |
|  | c. Explanation for lack of control group |  |  |  | + | + |  |  |
|  | d. Appropriateness of qualitative methodology |  |  |  |  |  |  | + |
|  | e. Appropriate Study design |  |  |  |  |  |  | ++ |
| 2. Managing bias in sampling or between groups | a. Sequence generation | ++ |  |  |  |  |  |  |
|  | b. Allocation concealment | ++ |  |  |  |  |  |  |
|  | c. Justification for sample choice |  |  |  | ++ | ++ |  |  |
|  | d. Intervention and control group selection designed to protect against systematic difference/selection bias |  | ++ |  |  |  |  |  |
|  | e. Comparability of groups |  |  |  |  |  | ++ |  |
|  | f. Sampling and recruitment |  |  |  |  |  |  | ++ |
| 3. Managing bias in outcome measurements and blinding | a. Blinding | ++ |  |  |  |  |  |  |
|  | b. Baseline measurement – protection against selection bias |  | ++ |  |  |  |  |  |
|  | c. Protection against contamination |  | ++ |  |  |  |  |  |
|  | d. Protection against secular changes |  |  | ++ |  |  |  |  |
|  | e. Protection against detection bias: blinded assessment of primary outcome measures | + | + | + | + | + | + |  |
|  | f. Reliable primary outcome measures | + | + | + | + | + | + | + |
|  | g. Comparability of outcomes |  |  |  |  |  | ++ |  |
| 4. Managing bias in follow up | a. Follow up of subjects (protection against exclusion bias) | + |  |  |  |  |  |  |
|  | b. Follow up of patients or episodes of care | + |  |  |  |  |  |  |
|  | c. Incomplete outcome data addressed | + | + | + | + | + | ++ | + |
| 5. Managing bias in other study aspects | a. Protection against detection bias: intervention unlikely to affect data collection | + | + | + | + | + |  |  |
|  | b. Protection against information bias |  |  |  |  |  | + |  |
|  | c. Data collection appropriate to address research aims |  |  |  |  |  |  | + |
|  | d. Attempts to mitigate effects of no control |  |  |  | ++ | ++ |  |  |
| 6. Analytical Rigour | a. Sufficient data points to enable reliable statistical inference |  |  | ++ |  |  |  |  |
|  | b. Shaping of intervention effect specified |  |  | + |  |  |  |  |
|  | c. Analysis sufficiently rigorous/free from bias | + | + | + | + | + | + | + |
| 7. Managing bias in reporting/ethical considerations | a. Free of selective outcome reporting | + | + | + | + | + | + | + |
|  | b. Limitations addressed | + | + | + | + | + | + | + |
|  | c. Conclusions clear and justified | + | + | + | + | + | + | + |
|  | d. Free of other bias | + | + | + | + | + | + | + |
|  | e. Ethics issues addressed | + | + | + | + | + | + | + |

<sup>a</sup>Applicability of quality criteria to each study design: + = Criteria to be included in quality assessment for study design; ++ = Mandatory criteria to be met in quality assessment; blank: criteria not to be applied in quality assessment for study design.
<sup>b</sup> Study designs: RCT: randomised controlled trial; CBA: controlled before-after; CITS: controlled interrupted time series; CS: cohort study; NCITS: non-controlled interrupted time series; NCBA: non-controlled before-after; QUAL: qualitative

**Supplementary Table 2:**
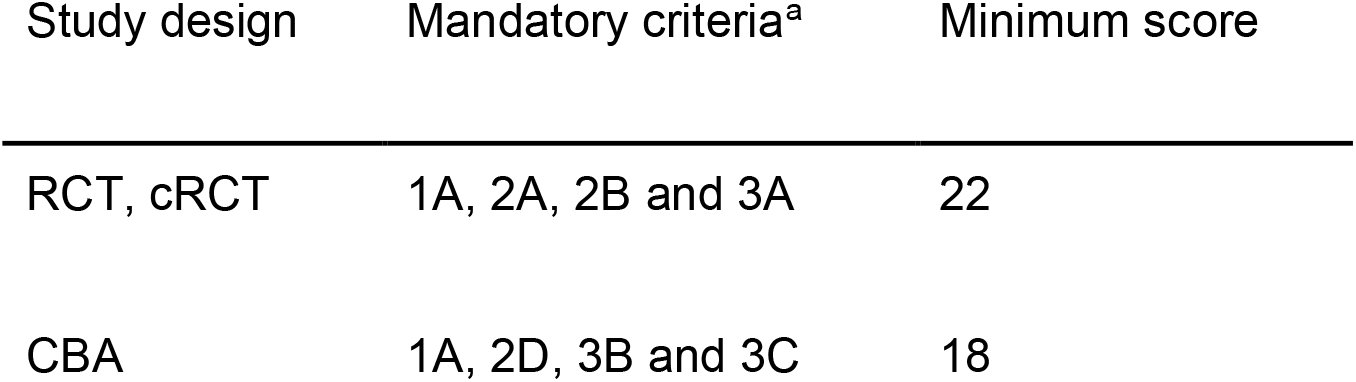
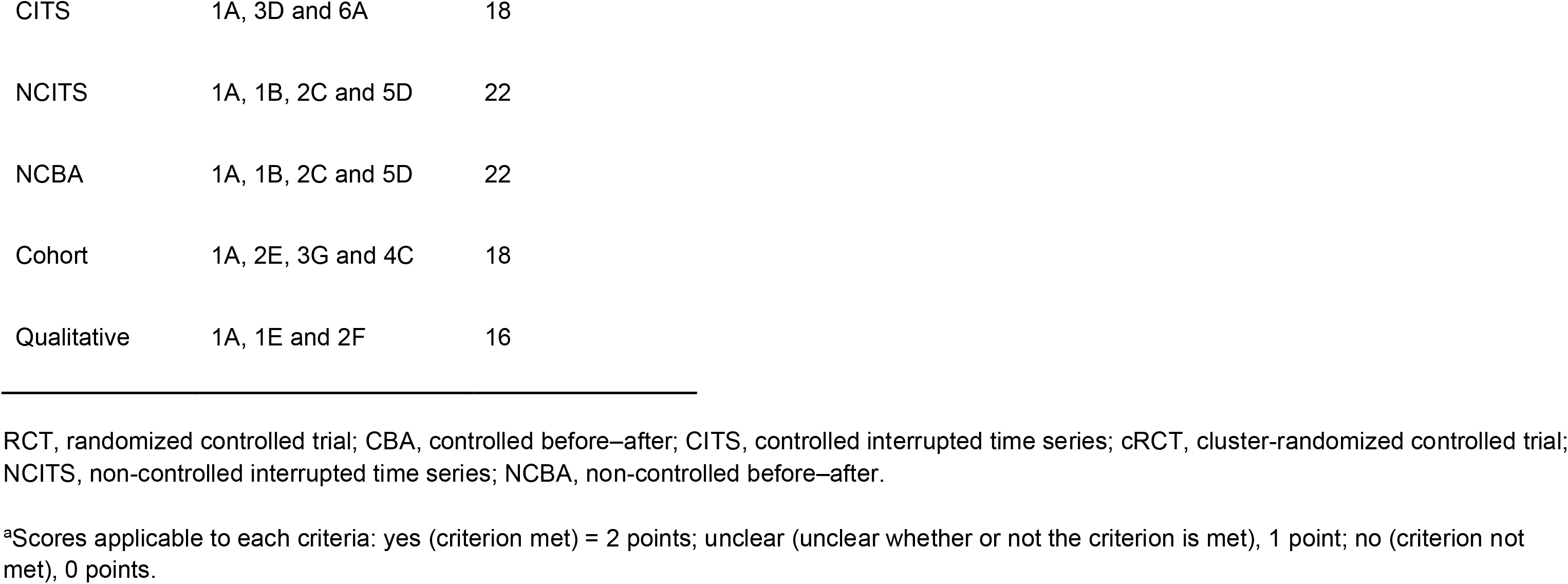
ICROMS Decision matrix: mandatory criteria and minimum score for study type to be included in review (from Zingg *et al*., 2016)^1^

